# Axonal damage and astrocytosis are biological correlates of grey matter network integrity loss: a cohort study in autosomal dominant Alzheimer disease

**DOI:** 10.1101/2023.03.21.23287468

**Authors:** L. Vermunt, C. Sutphen, E. Dicks, D.M. de Leeuw, R. Allegri, S.B. Berman, D.M. Cash, J.P. Chhatwal, C. Cruchaga, G. Day, M. Ewers, M. Farlow, N.C. Fox, B. Ghetti, N. Graff-Radford, J. Hassenstab, M. Jucker, C M. Karch, J. Kuhle, C. Laske, J. Levin, C.L Masters, E. McDade, H. Mori, J.C. Morris, R.J. Perrin, O. Preische, P.R. Schofield, M. Suárez-Calvet, C. Xiong, P. Scheltens, C.E. Teunissen, P.J. Visser, R.J. Bateman, T.L.S. Benzinger, A.M. Fagan, B.A. Gordon, B.M. Tijms, the Dominantly Inherited Alzheimer Network.

## Abstract

Brain development and maturation leads to grey matter networks that can be measured using magnetic resonance imaging. Network integrity is an indicator of information processing capacity which declines in neurodegenerative disorders such as Alzheimer disease (AD). The biological mechanisms causing this loss of network integrity remain unknown. Cerebrospinal fluid (CSF) protein biomarkers are available for studying diverse pathological mechanisms in humans and can provide insight into decline. We investigated the relationships between 10 CSF proteins and network integrity in mutation carriers (N=219) and noncarriers (N=136) of the Dominantly Inherited Alzheimer Network Observational study. Abnormalities in Aβ, Tau, synaptic (SNAP-25, neurogranin) and neuronal calcium-sensor protein (VILIP-1) preceded grey matter network disruptions by several years, while inflammation related (YKL-40) and axonal injury (NfL) abnormalities co-occurred and correlated with network integrity. This suggests that axonal loss and inflammation play a role in structural grey matter network changes.

**Key points:**

- Abnormal levels of fluid markers for neuronal damage and inflammatory processes in CSF are associated with grey matter network disruptions.
- The strongest association was with NfL, suggesting that axonal loss may contribute to disrupted network organization as observed in AD.
- Tracking biomarker trajectories over the disease course, changes in CSF biomarkers generally precede changes in brain networks by several years.

## INTRODUCTION

Brain areas implicated in similar functions show covariation in cortical morphology on magnetic resonance imaging (MRI), and these covariation patterns can be precisely quantified with a network approach [1-3]. In neurodegenerative diseases, such as Alzheimer disease (AD), grey matter networks become disrupted [3-9]. With increasing disease severity in AD, grey matter networks become more randomly organized, as indicated by a lower small world coefficient [9-11]. These network disruptions are related to impaired cognition and future cognitive decline, both in sporadic and autosomal dominant AD (ADAD) [10-16]. Network disruptions can already be detected in cognitively normal individuals with amyloid-β (Aβ) aggregation [10, 17, 18]. Still, the biological mechanisms that explain the deterioration of network organization remain unclear. Changes in grey matter networks could result from multiple pathophysiological processes such as synaptic dysfunction and loss, axonal degeneration, neuronal loss, and local swelling in response to infiltration of inflammatory cells. A better understanding of network disruptions over the course of AD may inform new hypotheses regarding what biological process maintain brain connectivity and preserve cognitive function.

In cerebrospinal fluid (CSF), proteins can be measured that reflect ongoing biological processes in the brain. CSF biomarkers are used for the biological definition of AD based on abnormal concentrations of Aβ (Aβ42/40 ratio), hyperphosphorylation of tau (181-phosphorylated fraction [pTau]), and neuronal injury (total tau [tTau]) [19]. In addition to these core AD measures, other biomarkers have robustly been related to AD, and provide information on additional pathological brain alterations occurring in the disease [20]: Increased levels of synaptosomal-associated protein-25 (SNAP-25) and neurogranin (Ng) levels are markers of pre-synaptic and post-synaptic dysfunction, respectively; visinin-like protein 1 (VILIP-1) a calcium-sensor protein increased with neuronal injury; and neurofilament light chain (NfL) of neuroaxonal damage [21-26]. In addition, chitinase-3-like protein 1 (YKL-40), an inflammation marker, and soluble TREM2 (sTREM2) [22, 23, 26-29] a marker of microglia, are also elevated in AD and provide insight into inflammatory processes. These markers have been associated with AD, but it remains unknown if they may influence grey matter networks.

We investigated this question in carriers of ADAD genetic mutations. This allows for investigation of the disease processes before onset of symptoms due to relatively conserved dementia onset age and few age-related co-pathologies [30]. We assessed the associations between both the core and emerging CSF biomarkers for AD and the individual grey matter network summary statistic, the ‘small world coefficient’ [11, 15, 16].We further tested within mutation carriers whether the relationships observed were dependent on disease stage, as measured by a combination of the pTau/Aβ_42_ ratio (normal, abnormal) and clinical dementia rating score (CDR).

## METHODS

### Participants and design

Data was obtained from the Dominantly Inherited Alzheimer Network Observational Study (DIAN-Obs) [31]. For the DIAN study, ADAD mutation carriers (MC) in the presenilin 1 [*PSEN1*], presenilin 2 [*PSEN2*] and amyloid precursor protein [*APP*] genes and their noncarrier (NC) family members undergo longitudinal clinical and cognitive examinations, neuroimaging and biospecimen donations. We evaluated data that passed quality control and was included in data freeze 12. Families with Flemish and Dutch mutations were excluded from analyses because these mutations result in a different phenotype, with primarily cerebral amyloid angiopathy. The study was approved by the ethical review board at Washington University, St. Louis, Missouri, USA and local IRBs. The estimated years to symptom onset (EYO) for each individual was defined as the mutation-specific mean age at onset subtracted from the individuals’ visit age (e.g. for the *PSEN1* G206A mutation, the mean age at onset is 53) [30]. Where the mutation age of onset was unknown, the family-specific parental age of disease onset was used instead. For example, if the mean age at symptom onset is 53 years for a specific family mutation, then a 43-year-old individual, regardless of mutation status, would have an EYO of -10. This indicates an individual with the mutation is expected to show clinical symptoms of AD 10 years later and allows comparison of biomarkers with the NCs on the same timeline, as well as between MCs and NCs from different families and mutations. For the biomarker comparisons, we selected the first visit at which individuals had both CSF and MRI data available.

### Group definitions

Participants were stratified in two ways. The first set of analyses focused on comparing all MCs to their familial NC controls. In the second set of analyses, MCs were split into 4 disease stage groups based upon their biomarker status and clinical dementia rating (CDR) [32]. Group 1 had a normal CSF ratio of pTau/Aβ_42_ (< 0.019 [33]) (indicating absence of underlying brain amyloid). Group 2-4 had abnormal ratios (indicating presence of amyloid) and increasing CDRs of: group 2: CDR= 0, no impairment; group 3: CDR = 0.5, very mild dementia; group 4: CDR >= 1 mild to severe dementia.

### MR preprocessing

MR scans were collected and preprocessed according the protocols of the Alzheimer’s Disease Neuroimaging Initiative (ADNI)(1.1□by□1.1□by□1.2 mm^3^ voxels, repetition time = 2300, echo time = 2.95, flip angle 9°), described in detail in [34-36]. For the network extractions, T1-weighted scans were first segmented into grey matter, white matter and CSF in native space with Statistical Parametric Mapping 12 (SPM12; Wellcome Trust Centre for Neuroimaging, UCL Institute of Neurology, London, UK). The segmentations were checked and resliced into 2mm by 2mm by 2mm voxels, and this was the input for the grey matter network extraction.

### Calculation of grey matter network metrics

Single-subject grey matter network metrics were extracted from preprocessed grey matter segmentations according to previously published procedures (https://github.com/bettytijms/Single_Subject_Grey_Matter_Networks)[2] as follows: Grey matter segmentations were parcellated into cubes of 3 by 3 by 3 voxels, which formed the nodes of the network. The Pearson’s correlation coefficient was then calculated for grey matter intensities across the voxels for each pair of cubes. Next, correlation values were binarized by retaining only significant connections after correction for multiple comparisons with a permutation based method. Finally, we calculated for each network the small world coefficient using scripts from the brain connectivity toolbox (https://sites.google.com/site/bctnet/ [37] modified for large sized networks. In this study, we use the small world coefficient, which is a whole brain summary statistic and normalized for individual differences in degree and size of networks. The small world coefficient is the result of dividing the clustering coefficient by path length. Values of small world range between 0 and infinity with values near 1 indicating a random organization. The most efficient network organization is characterized by a small world property with a high level of clustering combined with a relatively short path length. This will result in a small world index higher than 1. [38, 39], indicating efficient information processing.

### Cerebrospinal fluid markers

Participants underwent lumbar puncture (LP) after overnight fasting. Samples were collected via gravity drip in polypropylene tubes and sent on dry ice to the DIAN biomarker laboratory at Washington University. The samples were then thawed and aliquoted (0.5mL) in polypropylene tubes, stored at -84°C before measurements of SNAP-25, Ng, VILIP-1 and YKL-40. Additional aliquots of each sample were shipped on dry ice for the measurements of Aβ_40_, Aβ_42_, pTau and tTau by the Shaw laboratory at the University of Pennsylvania [40], of NfL by the Kuhle laboratory in Basel [41], and of sTREM2 by the Haass laboratory in Münich [29, 42]. For details on the protocols, see [24, 25, 27, 43]. Briefly, Aβ_40_, Aβ_42_, pTau and tTau levels were determined using the automated Elecsys assay, and values of Aβ_40_ and Aβ_42_ outside the measurement ranges were extrapolated on the calibration curve [43]. SNAP-25, Ng and VILIP-1 were measured with antibodies developed in the laboratory of Dr. Jack Ladenson at Washington University in St. Louis, as part of micro-particle-based immunoassays using the Singulex (now part of EMD Millipore; Alameda, CA) Erenna system [24]. YKL-40 was measured with plate-based enzyme-linked immunoassay (MicroVue ELISA; Quidel, San Diego, CA) [24]. NfL was measured on a single-molecule array (SIMOA) using the capture monoclonal antibody 47:3 and biotinylated detection antibody 2:1 (UmanDiagnostics AB, Sweden) [25]. Soluble TREM2 (sTREM2) was measured using the Meso Scale Discovery (MSD) platform with an in-house developed ELISA based on commercially available antibodies [27, 44]. The sTREM2 concentrations are reported relative to an internal standard sample that was loaded onto each assay plate as a way to account for inter-plate variation.

### Statistical analysis

In all linear models tTau, pTau, SNAP-25, Ng, VILIP-1, YKL-40, NfL and sTREM2 were log-transformed to approach normality. To aid comparability of slope estimates, the variables were Z-transformed according to the entire cohort. We tested the associations between the CSF biomarkers as predictors and the small world coefficient as the outcome with three linear regression models. Model 1 was adjusted for sex; Model 2 also included a fixed term for mutation status and its interaction with the predictor; Model 3 had additional adjustment for age effects as main effect. We further performed a subgroup analysis *within* MCs only so to investigate disease stage effects: these models included the CSF biomarkers as predictor, a fixed term for the interaction between the severity groups and their predictor. In cases of significant interactions terms, we ran post-hoc pairwise comparisons using the Tukey HSD procedure.

Lastly, we estimated trajectories for all markers studied by EYO, using a previously developed Bayesian inference linear mixed effect model [35, 45] to obtain insight into the relative ordering of biomarker trajectories (details in Supplement). Before fitting this model, the CSF and MRI biomarkers were Z-scored to young NCs (<40 years old, n=81, table 1). All statistical analyses were conducted in R (version 3.5.3) using the emmeans. car, lmer, rstan and rstanarm-packages [46].

**Table 1.** Demographics and baseline summary data on predictors and outcomes.

|  | Noncarriers (NCs) |  | Mutation carriers (MCs) |  |  |  |  |
| --- | --- | --- | --- | --- | --- | --- | --- |
|  | All<br>(n=136) | NCs < 40<br>years old<br>(n=81) | All<br>(n=216) | MCs: ratio<br>neg (n=84) | MCs: CDR<br>0, ratio pos<br>(n=63) | MCs: CDR<br>0.5, ratio<br>pos (n=43) | MCs: CDR<br>1-3, ratio<br>pos (n=26) |
| **Demographics** |  |  |  |  |  |  |  |
| **N (%) Male** | 53 (39%) | 32 (40%) | 96 (44%) | 35 (42%) | 28 (44%) | 19 (44%) | 14 (54%) |
| **Age, years** | 38 ± 12 | 31 ± 6 | 39 ± 10 | 32 ± 8 | 38 ± 9 | 47 ± 9 | 47 ± 9 |
| **EYO, years** | -10 ± 12 | -17 ± 9 | -9 ± 11 | -17 ± 8 | -8 ± 7 | 1 ± 6 | 4 ± 4 |
| **Years of education, median±IQR** | 15 ± 3 | 15 ± 2 | 14 ± 4 | 15 ± 3 | 15 ± 4 | 14 ± 4 | 12 ± 2 |
| **CDR (0/0.5-1/2-3), N** | 131/5/0 | 0/2/0 | 142/66/8 | 79/5/0 | 63/0/0 | 0/43/0 | 0/18/8 |
| **MMSE, median±IQR** | 30 ± 1 | 30 ± 1 | 29 ± 3 | 29 ± 1 | 29 ± 2 | 26 ± 4 | 16 ± 10 |
| **Grey matter network** |  |  |  |  |  |  |  |
| **Small world coefficient** | 1.62 ± 0.05 | 1.65 ± 0.05 | 1.59 ± 0.08 | 1.64 ± 0.06 | 1.60 ± 0.05 | 1.55 ± 0.08 | 1.46 ± 0.07 |
| **Traditional CSF markers** |  |  |  |  |  |  |  |
| **Aβ<sub>42</sub> pg/ml** | 1,407 ± 466 | 1,292 ± 442 | 974 ± 634 | 1,526 ± 655 | 716 ± 279 | 553 ± 208 | 510 ± 217 |
| **Aβ<sub>40</sub> pg/ml** | 15,698 ± 4418 | 14,398 ± 4,204 | 14,862 ± 4760 | 15,607 ± 5,080 | 15,004 ± 4,749 | 14,483 ± 4,114 | 12,741 ± 4,241 |
| **pTau pg/ml** | 14 ± 5 | 13 ± 4 | 31 ± 23 | 14 ± 4 | 32 ± 18 | 46 ± 23 | 57 ± 28 |
| **tTau pg/ml** | 169 ± 55 | 154 ± 49 | 290 ± 162 | 177 ± 48 | 305 ± 120 | 375 ± 142 | 475 ± 241 |
| **Ratio aβ<sub>42/40</sub>** | 0.089 ± 0.01 | 0.089 ± 0.007 | 0.066 ± 0.035 | 0.099 ± 0.031 | 0.049 ± 0.017 | 0.039 ± 0.012 | 0.042 ± 0.015 |
| **ratio Aβ<sub>42/40</sub> ↓0.075, N (%)** | 6 (4%) | 1 (1%) | 144 (67%) | 19 (23%) | 58 (92%) | 42 (98%) | 25 (96%) |
| **ratio pTau/ Aβ<sub>42</sub>** | 0.010 ± 0.004 | 0.010 ± 0.002 | 0.052 ± 0.053 | 0.010 ± 0.004 | 0.051 ± 0.034 | 0.091 ± 0.049 | 0.120 ± 0.065 |
| **ratio pTau/Aβ<sub>42</sub> ↑0.0198, N (%)** | 2 (1%) | 0 (0%) | 132 (61%) | - | - | - | - |
| **Emerging CSF markers** |  |  |  |  |  |  |  |
| **SNAP-25 pg/ml** | 3.6 ± 1.3 | 3.2 ± 1.1 | 4.6 ± 1.9 | 3.6 ± 1.1 | 4.5 ± 1.5 | 5.2 ± 1.7 | 6.4 ± 2.7 |
| **Ng pg/ml** | 1,563 ± 741 | 1,447 ± 765 | 2,297 ± 1,212 | 1,638 ± 682 | 2,526 ± 1,109 | 2,673 ± 1,164 | 3,120 ± 1,748 |
| **NfL pg/ml** | 793 ± 544 | 564 ± 396 | 1,939 ± 1,762 | 531 ± 190 | 1,033 ± 650 | 2,630 ± 1,643 | 3,873 ± 1,657 |
| **VILIP-1 pg/ml** | 133 ± 50 | 122 ± 48 | 174 ± 79 | 135 ± 47 | 179 ± 71 | 198 ± 75 | 236 ± 114 |
| **YKL-40 ng/ml** | 133 ± 66 | 98 ± 37 | 173 ± 88 | 109 ± 37 | 169 ± 69 | 229 ± 81 | 280 ± 89 |
| **sTREM2, relative to reference sample** | 0.47 ± 0.22 | 0.43 ± 0.21 | 0.58 ± 0.29 | 0.42 ± 0.15 | 0.48 ± 0.28 | 0.73 ± 0.3 | 0.74 ± 0.3 |

Legend: CSF biomarkers not available for the whole sample: SNAP (n=330), Ng (n=331), VILIP1 (n=330), YKL40 (n=331), NfL (n=169 [incl. 19 with no MRI data], sTREM2 (n=164).

## RESULTS

The presented analyses included 219 MCs and 136 NCs (age mean±SD 39±11; EYO mean±SD -9±11). In the MC group, 84 (39%) individuals had normal CSF ratio pTau/Aβ_42_. Among MCs with an abnormal CSF pTau/Aβ_42_ ratio, 63 (29%) individuals had CDR 0, 43 (20%) CDR 0.5 and 26 (12%) CDR 1-3. The group characteristics are shown in Table 1.

### Associations between CSF biomarkers and the small world coefficient

Across the whole group, we found that all AD markers were related to alterations in grey matter networks (Table 2). Higher levels of NfL were most strongly related to lower small world values (β = -0.72 [CI 95% - 0.83, -0.61]; p<0.001), followed by YKL-40 (β = -0.53 [CI 95% -0.63, -0.44]; p<0.001), and pTau (β = -0.53 [CI 95% -0.61, -0.44]; p<0.001, Table 2). Models taking into account interaction terms for mutation status and CSF predictor were significant for SNAP-25, Ng, pTau, tTau, NfL, VILIP-1 and YKL-40 (p<0.05, Fig. 1). Post-hoc comparisons showed that higher levels of SNAP-25 (−0.37 [CI 95%, -0.50,-0.24]) and Ng (−0.35 [CI 95%, -0.48,-0.21]), pTau (−0.58 [CI 95%, -0.69,-0.48]), tTau (−0.55 [CI 95%, -0.67,-0.44]) and VILIP-1 (−0.29 [CI 95%, -0.42,-0.16]) were related to lower small world values specifically in MCs. The association of higher NfL and YKL-40 and lower small world values was observed in both MCs and NCs, and this was stronger in MCs for YKL-40 (NfL: MC = -0.76 [CI 95%, -0.89, -0.64] & NC = -0.44 [CI 95%, - 0.77,-0.17]; YKL-40: MC = -0.61 [CI 95%, -0.72, -0.49] & NC = -0.32 [CI 95%, -0.48,-0.17]). When repeating models correcting for age, interaction effects for mutation status changed by 0.14-0.39 and remained significant for SNAP-25, Ng, pTau, tTau, NfL and YKL-40 (p<0.05), but evidence for VILIP-1 is weaker (p=0.08). Next, we studied in MCs whether the observed associations were specific to disease stage (Table S1, Fig. 2). No significant interaction terms with disease stage were observed, suggesting that associations of biomarkers and small world values were not specific to a certain stage.

**Table 2.**
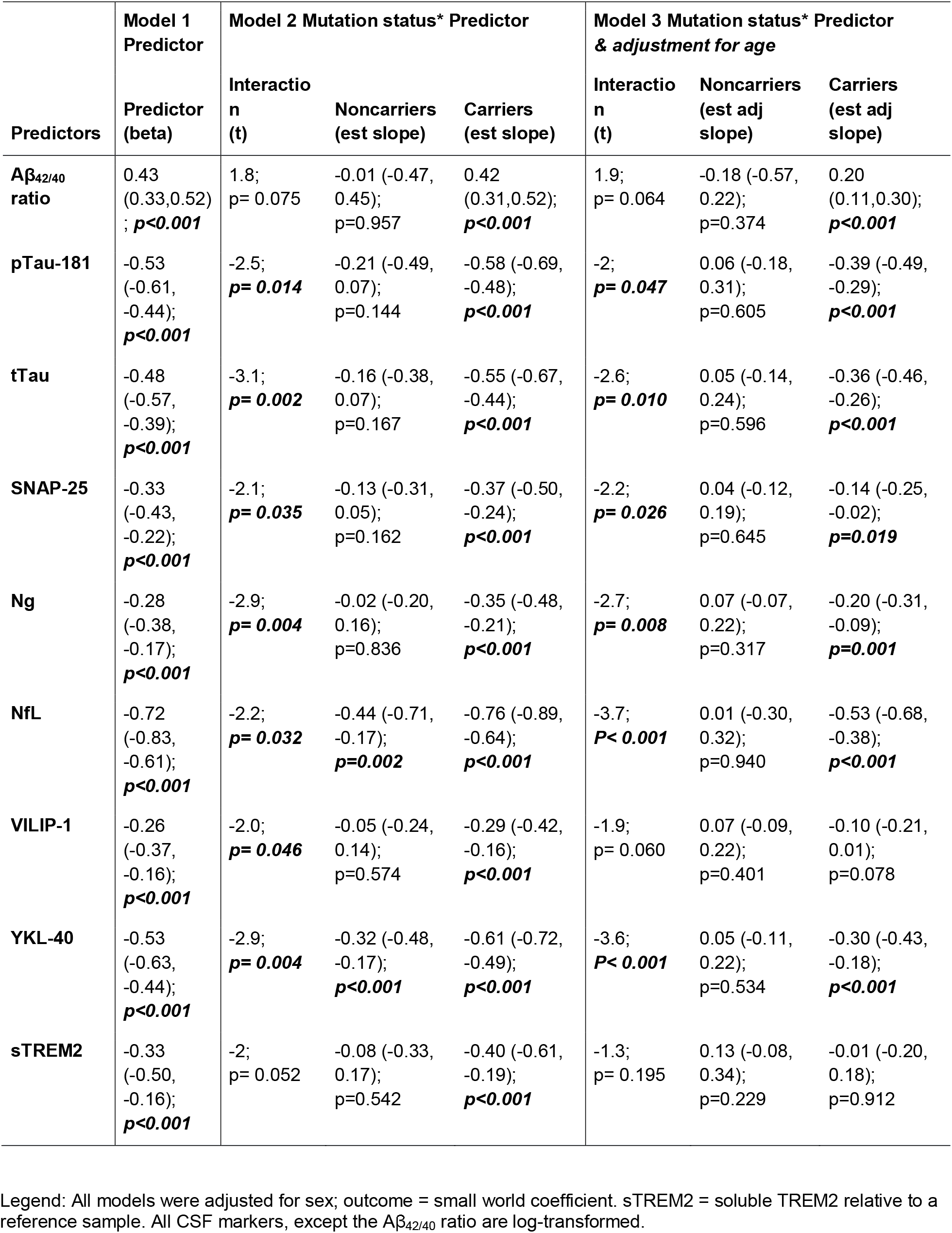
Associations between CSF markers and the small world coefficient

| Predictors | Model 1<br>Predictor | Model 2 Mutation status* Predictor |  |  | Model 3 Mutation status* Predictor<br>& adjustment for age |  |  |
| --- | --- | --- | --- | --- | --- | --- | --- |
|  | Predictor<br>(beta) | Interactio<br>n<br>(t) | Noncarriers<br>(est slope) | Carriers<br>(est slope) | Interactio<br>n<br>(t) | Noncarriers<br>(est adj<br>slope) | Carriers<br>(est adj<br>slope) |
| **Aβ<sub>42/40</sub>**<br>**ratio** | 0.43<br>(0.33,0.52)<br>; **p<0.001** | 1.8;<br>p= 0.075 | -0.01 (-0.47,<br>0.45);<br>p=0.957 | 0.42<br>(0.31,0.52);<br>**p<0.001** | 1.9;<br>p= 0.064 | -0.18 (-0.57,<br>0.22);<br>p=0.374 | 0.20<br>(0.11,0.30);<br>**p<0.001** |
| **pTau-181** | -0.53<br>(-0.61,<br>-0.44);<br>**p<0.001** | -2.5;<br>**p= 0.014** | -0.21 (-0.49,<br>0.07);<br>p=0.144 | -0.58 (-0.69,<br>-0.48);<br>**p<0.001** | -2;<br>**p= 0.047** | 0.06 (-0.18,<br>0.31);<br>p=0.605 | -0.39 (-0.49,<br>-0.29);<br>**p<0.001** |
| **tTau** | -0.48<br>(-0.57,<br>-0.39);<br>**p<0.001** | -3.1;<br>**p= 0.002** | -0.16 (-0.38,<br>0.07);<br>p=0.167 | -0.55 (-0.67,<br>-0.44);<br>**p<0.001** | -2.6;<br>**p= 0.010** | 0.05 (-0.14,<br>0.24);<br>p=0.596 | -0.36 (-0.46,<br>-0.26);<br>**p<0.001** |
| **SNAP-25** | -0.33<br>(-0.43,<br>-0.22);<br>**p<0.001** | -2.1;<br>**p= 0.035** | -0.13 (-0.31,<br>0.05);<br>p=0.162 | -0.37 (-0.50,<br>-0.24);<br>**p<0.001** | -2.2;<br>**p= 0.026** | 0.04 (-0.12,<br>0.19);<br>p=0.645 | -0.14 (-0.25,<br>-0.02);<br>**p=0.019** |
| **Ng** | -0.28<br>(-0.38,<br>-0.17);<br>**p<0.001** | -2.9;<br>**p= 0.004** | -0.02 (-0.20,<br>0.16);<br>p=0.836 | -0.35 (-0.48,<br>-0.21);<br>**p<0.001** | -2.7;<br>**p= 0.008** | 0.07 (-0.07,<br>0.22);<br>p=0.317 | -0.20 (-0.31,<br>-0.09);<br>**p=0.001** |
| **NfL** | -0.72<br>(-0.83,<br>-0.61);<br>**p<0.001** | -2.2;<br>**p= 0.032** | -0.44 (-0.71,<br>-0.17);<br>**p=0.002** | -0.76 (-0.89,<br>-0.64);<br>**p<0.001** | -3.7;<br>**P< 0.001** | 0.01 (-0.30,<br>0.32);<br>p=0.940 | -0.53 (-0.68,<br>-0.38);<br>**p<0.001** |
| **VILIP-1** | -0.26<br>(-0.37,<br>-0.16);<br>**p<0.001** | -2.0;<br>**p= 0.046** | -0.05 (-0.24,<br>0.14);<br>p=0.574 | -0.29 (-0.42,<br>-0.16);<br>**p<0.001** | -1.9;<br>p= 0.060 | 0.07 (-0.09,<br>0.22);<br>p=0.401 | -0.10 (-0.21,<br>0.01);<br>p=0.078 |
| **YKL-40** | -0.53<br>(-0.63,<br>-0.44);<br>**p<0.001** | -2.9;<br>**p= 0.004** | -0.32 (-0.48,<br>-0.17);<br>**p<0.001** | -0.61 (-0.72,<br>-0.49);<br>**p<0.001** | -3.6;<br>**P< 0.001** | 0.05 (-0.11,<br>0.22);<br>p=0.534 | -0.30 (-0.43,<br>-0.18);<br>**p<0.001** |
| **sTREM2** | -0.33<br>(-0.50,<br>-0.16);<br>**p<0.001** | -2;<br>p= 0.052 | -0.08 (-0.33,<br>0.17);<br>p=0.542 | -0.40 (-0.61,<br>-0.19);<br>**p<0.001** | -1.3;<br>p= 0.195 | 0.13 (-0.08,<br>0.34);<br>p=0.229 | -0.01 (-0.20,<br>0.18);<br>p=0.912 |

Legend: All models were adjusted for sex; outcome = small world coefficient. sTREM2 = soluble TREM2 relative to a reference sample. All CSF markers, except the Aβ <sub>42/40</sub> ratio are log-transformed.

**Figure 1.**
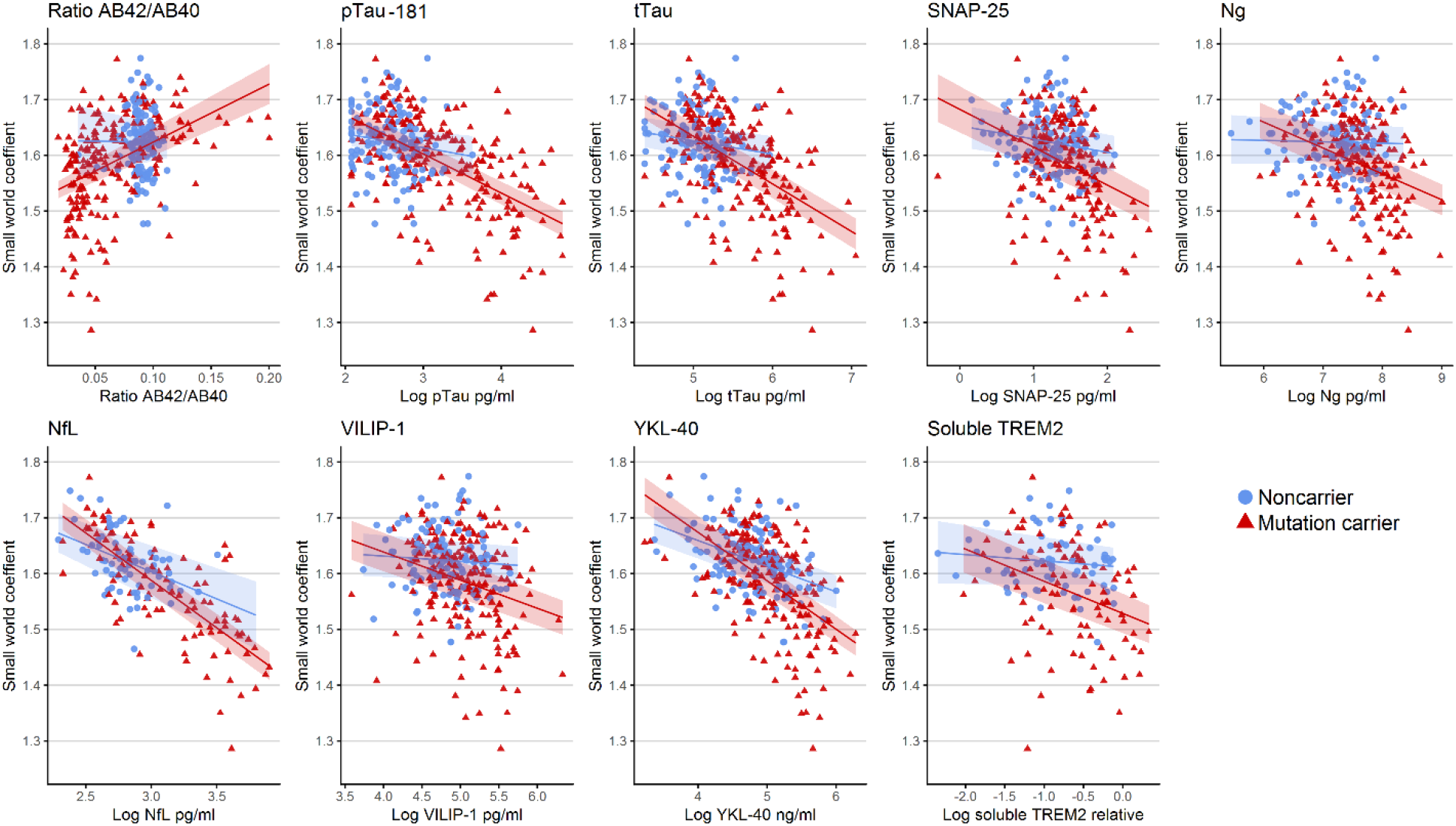
Associations between CSF biomarkers and grey matter networks for mutation carriers and noncarriers. Legend: Adjusted for sex. Prediction with 95% confidence intervals. sTREM2 = soluble TREM2 relative to a reference sample.

**Figure 2.**
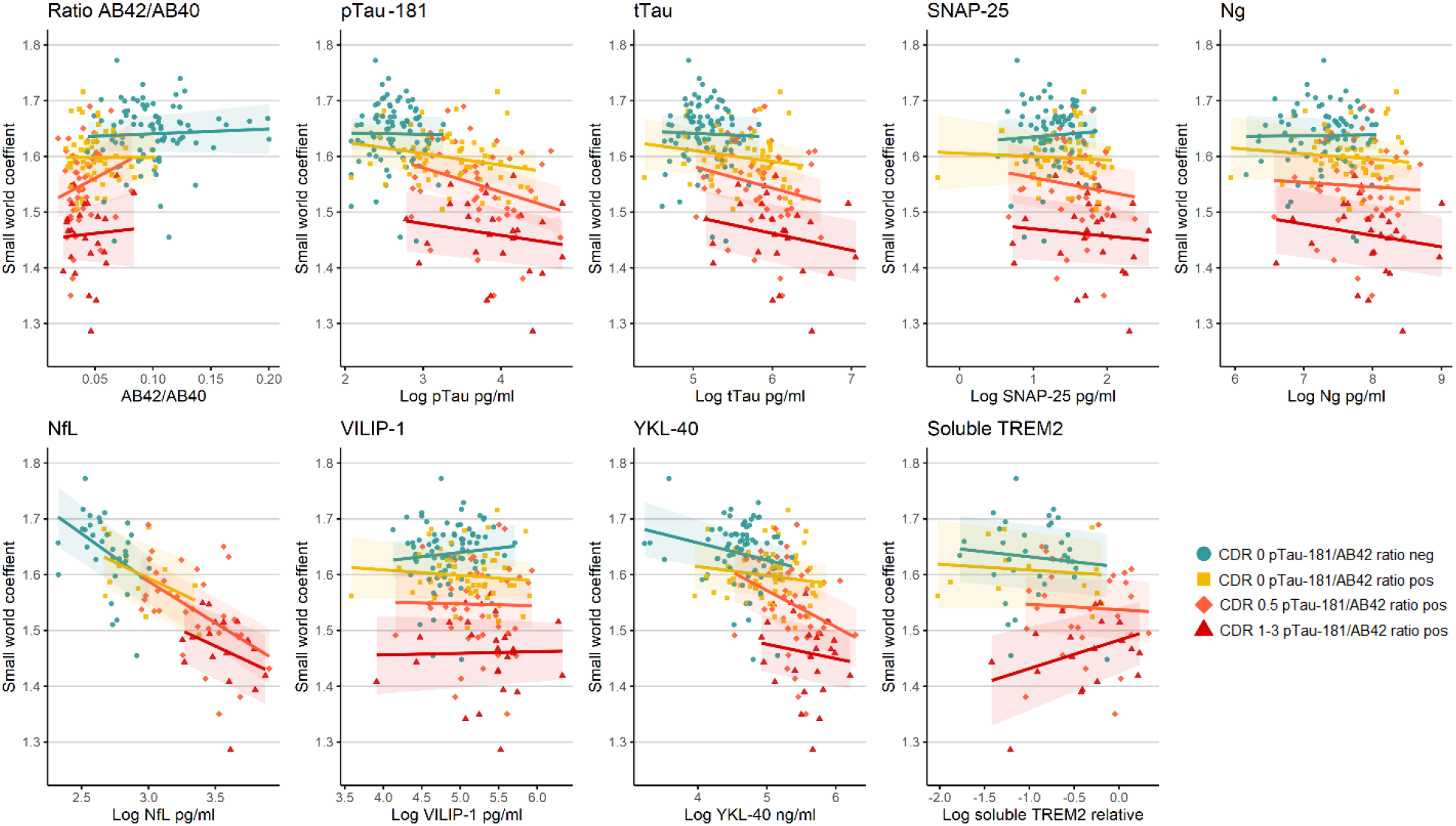
Associations between CSF biomarkers and grey matter networks within mutation carriers by disease stage. Legend: Adjusted for sex. Prediction with 95% confidence intervals. sTREM2 = soluble TREM2 relative to a reference sample.

### Small world coefficient and CSF biomarker trajectory by EYO

Finally, we estimated trajectories for all CSF and structural MRI markers according to EYO for the MCs, NCs, and the difference between MCs and NCs (Fig. S3; Table S2 and S3). Biomarker trajectories of the Aβ_42/40_ ratio, pTau, tTau, SNAP-25, Ng, and VILIP-1 levels were different in MCs as compared to NCs before differences were observed in grey matter networks. NfL and YKL-40 trajectories were abnormal around the same time as small-world values, and sTREM2 and Aβ_40_ showed abnormal levels in MCs compared to NCs later than small-world values.

## DISCUSSION

The main finding of our study is that CSF pathologic biomarkers showed associations with alterations in grey matter networks, and that axonal damage as measured with NfL showed the strongest relationship with worse grey matter network disruptions. Increased concentrations of the CSF markers for hyperphosphorylation of tau (pTau), neuronal injury and death (tTau and VILIP-1), and specific synaptic injury (SNAP-25 and Ng) were related to worse grey matter network organization in the MCs only. The observed associations were not dependent on staging based on a combination of the pTau/Aβ_42_ ratio and the global CDR, suggesting that they were similar across disease development. Most CSF biomarkers showed abnormal levels before grey matter network abnormality in the MCs compared to the NCs, and for NfL and YKL-40 the timing was closest aligned to grey matter network alterations.

To date, only the role of Aβ aggregation and tau had been studied in relation to grey matter network integrity in individuals with AD [17, 18, 47]. Those findings suggested that grey matter networks are sensitive to brain structural changes related to amyloid and tau aggregation in sporadic AD. Here, we found also that lower Aβ_42/40_ ratios were associated with grey matter network disruptions. We further detected relationships between markers of other pathological processes in ADAD and grey matter network disruptions. The most pronounced association was observed for NfL, which suggests that loss of axonal integrity plays a critical role in the loss of grey matter network organization. The link between axonal tract damage and deterioration of grey matter covariance in AD supports the idea that grey matter covariance networks reflect, at least in part, axonal connectivity.

We also observed that higher levels of the synaptic markers (SNAP-25 and Ng), pTau and neuronal damage (tTau and VILIP-1) were associated with grey matter network disruption, and this was specific for MCs. Synaptic maturation and co-activation are associated with grey matter covariance during development, promoting brain connectivity [3]. In AD, synaptic damage in neurodegeneration could possibly influence brain connectivity in the opposite way. A study by Pereira and colleagues showed that synaptic markers like SNAP-25 and Ng were associated with worse functional connectivity in the default mode network and worse memory which supports this hypothesis [48]. The biomarker trajectories suggest that synaptic damage and neuronal loss precedes the changes we observe with MRI, which could be a downstream effect. Recent analyses had already demonstrated that CSF pTau and tTau increase very early in the course of ADAD, in a more parallel fashion with amyloid aggregation than according to hypothetical models [33, 49]. Our findings suggest that loss of connectivity structures at the microscale (in neurons), could lead to disrupted whole-brain connectivity as measured on MRI. Longitudinal studies are needed to further examine the temporal relationship of these processes in more detail.

The associations between increased NfL and YKL-40, and network disruptions were also observed in NCs which suggests an effect of aging or another non-AD process. Previous studies have shown that during aging NfL and YKL-40 levels increase and grey matter network measures decline, additive to elevations in predementia AD (Chen, He et al. 2011, Alcolea, Vilaplana et al. 2015, Sutphen, Jasielec et al. 2015, Bridel, van Wieringen et al. 2019, Preische, Schultz et al. 2019, Schindler, Li et al. 2019, Luckett, Chen et al. 2022, Meeker, Butt et al. 2022). Our finding suggests that also in non-AD related aging, loss of axonal integrity and inflammation may affect grey matter network integrity. Therefore, the next step would be to investigate to what extent these changes in axonal integrity and inflammation contribute to the vulnerability of the brain to neurodegeneration and how it reflects cognitive decline in normal and non-AD related aging of the brain. sTREM2, released by microglia, fluctuates over the course of AD, with an increase close to symptom onset [27, 28, 50]. In the current study, the association between sTREM2 levels and grey matter networks disappeared in mutation carriers when analyses were corrected for age. We did not find compelling evidence that inflammatory processes due to microglial activity, as reflected by sTREM2 increases, may not be linearly related to grey matter morphological change.

A strength of the present study is that we evaluated the pathophysiology over the full course of AD. Investigating MCs from the DIAN study, along with NCs, is a powerful approach to investigate multiple disease processes that may contribute to grey matter network disruption. Due to the causative genes in dominantly inherited AD, the cross-sectional trajectory can inform longitudinal changes. Still, the reality is more complex [51], meaning further study in a longitudinal design is needed to understand the drivers and downstream effects in disease progression. A shortcoming of fitting AD biomarker trajectories over the estimated years to symptom onset is that results in part depend on sample sizes and model assumptions. All EYOs of divergence were similar to previous studies, except for Ng and YKL-40, which is an indication of the level of robustness across modeling methods [26]. sTREM2 was estimated to change later compared to a recent large study, but consistent with an older study with the same data [27, 29]. Of note regarding generalization of biomarker trajectories, estimated with cross-sectional data often diverse earlier compared to the estimates based on longitudinal data as there is a strong bias towards earlier years from cross-sectional data [51]. In addition, the exact meaning of biomarker levels is not fully understood, as we were unable to investigate brain tissue as part of this study. Another limitation is that we assessed linear relationships between CSF and grey matter network values, which may underestimate existing relationships. Therefore, we evaluated whether patterns depended on disease severity, which may give rise to non-linear patterns. Still, some of those disease stage groups were of small size, and larger samples are required to further investigate these relationships in detail. Lastly, in this study we studied a primary summary measure of network organization in order to reduce the number of comparisons and increase the interpretability of the data. The findings warrant follow-up research to evaluate whether associations are specific for specific brain areas or network measures and their generalization to sporadic (late onset) AD.

To summarize, loss of synaptic integrity and, in particular, axonal integrity, as evidenced by increased NfL in CSF, appears to be related to disrupted grey matter network organization in ADAD. These findings suggest that normalization of neuronal injury or synaptic processes might lead to stabilization or improvement of grey matter network integrity.

## Data Availability

The data set can be requested by qualified researchers through the DIAN website, see for details: https://dian.wustl.edu/our-research/for-investigators/dian-observational-study-investigator-resources/data-request-form/

## Acknowledgements

Data collection and sharing for this project was supported by The Dominantly Inherited Alzheimer’s Network (DIAN, UF1AG032438) funded by the National Institute on Aging (NIA), the German Center for Neurodegenerative Diseases (DZNE), Raul Carrea Institute for Neurological Research (FLENI), Partial support by the Research and Development Grants for Dementia from Japan Agency for Medical Research and Development, AMED (17929884, 16815631), and the Korea Health Technology R&D Project through the Korea Health Industry Development Institute (KHIDI).This manuscript has been reviewed by DIAN Study investigators for scientific content and consistency of data interpretation with previous DIAN Study publications. We acknowledge the altruism of the participants and their families and contributions of the DIAN research and support staff at each of the participating sites for their contributions to this study.

Further support for the study by Alzheimer Nederland Fellowship 2018 (L.V.), Willman Scholar Fund of the Barnes Jewish Hospital Foundation K01 AG053474 (B.A.G).

Computations were performed using the facilities of the Washington University Center for High Performance Computing, which were partially funded by NIH grants 1S10RR022984–01A1 and 1S10OD018091–01.

## Supplemental methods and materials

**Table S1.** Association of CSF markers with the small world coefficient within MCs by severity groups

| Predictors | Within MCs: interaction of group and CSF predictor |  |  |  |  |
| --- | --- | --- | --- | --- | --- |
|  | Interaction (F) | Ratio negative (est slope) | Ratio positive & CDR 0 (est slope) | Ratio positive & CDR 0.5 (est slope) | Ratio positive & CDR 1-3 (est slope) |
| **Aβ<sub>42/40</sub> ratio** | 0.63; p=0.59 | 0.03 (-0.13,0.20); p=0.68 | 0.0 (-0.36,0.35); p>0.99 | 0.44 (-0.14,1.02); p=0.13 | 0.09 (-0.53,0.71); p=0.77 |
| **pTau** | 0.68; p=0.57 | -0.02 (-0.41,0.37); p=0.92 | -0.17 (-0.4,0.05); p=0.13 | **-0.36 (-0.67,-0.06); p=0.02** | -0.18 (-0.54,0.19); p=0.34 |
| **tTau** | 0.34; p=0.8 | -0.04 (-0.36,0.27); p=0.78 | -0.13 (-0.35,0.1); p=0.27 | -0.25 (-0.56,0.05); p=0.10 | -0.2 (-0.52,0.12); p=0.23 |
| **SNAP-25** | 0.36; p=0.78 | 0.06 (-0.19,0.31); p=0.64 | -0.03 (-0.24,0.17); p=0.75 | -0.14 (-0.43,0.16); p=0.36 | -0.07 (-0.34,0.2); p=0.61 |
| **Neurogranin** | 0.24; p=0.87 | 0.01 (-0.21,0.23); p=0.92 | -0.07 (-0.3,0.16); p=0.53 | -0.06 (-0.36,0.24); p=0.69 | -0.15 (-0.47,0.17); p=0.35 |
| **NfL** | 0.16; p=0.92 | **-0.77 (-1.36,-0.18); p=0.01** | -0.51 (-1.17,0.14); p=0.12 | **-0.67 (-1.04,-0.3); p<0.01** | -0.50 (-1.26,0.26); p=0.2 |
| **VILIP-1** | 0.30; p=0.83 | 0.09 (-0.15,0.33); p=0.46 | -0.06 (-0.26,0.15); p=0.58 | -0.02 (-0.3,0.26); p=0.89 | 0.02 (-0.25,0.28); p=0.9 |
| **YKL-40** | 0.75; p=0.52 | -0.22 (-0.45,0.02); p=0.07 | -0.11 (-0.35,0.13); p=0.37 | **-0.46 (-0.84,-0.07); p=0.02** | -0.18 (-0.66,0.3); p=0.46 |
| **sTREM2** | 0.82 ; p=0.48 | -0.13 (-0.57,0.32); p=0.58 | -0.07 (-0.5,0.36); p=0.74 | -0.07 (-0.52,0.38); p=0.76 | 0.36 (-0.15,0.86); p=0.17 |

Legend: All models were adjusted for sex; outcome = small world coefficient. sTREM2 = soluble TREM2 relative to a reference sample. All CSF markers, except the ratio, are log-transformed.

### Details statistical methods of biomarker trajectory model

The statistical model to fit biomarker trajectories by EYO (Gordon, Blazey et al. 2018 Lancet Neurology) accounted for non-linear effects by using a restricted cubic spline to model EYO, with knots on the 0.1, 0.5 and 0.9 of the distribution. The models had fixed terms for EYO, mutation status, their interaction and a random effect for family cluster. Models were adjusted for sex and for the small world coefficient additionally for total grey matter volume. For the trajectories we used the biomarker data of the first available visit (Table S1 below). Model parameters were estimated with Hamiltonian Markov chain Monte Carlo sampling of the posterior distribution, with cauchy prior, 10,000 iterations in 8 chains, and thinning of 10 in the STAN and rstanarm package for R (Carpenter, Gelman et al. 2017). The EYO point of divergence is when the 99% credible intervals of the difference distribution between MCs and NCs did not overlap 0. We also provide the 95% and 99.5% of the credible intervals (Table S2).

R code: model_fit <-stan_glmer(standardized_biomarker_value ∼ (1 | family_id) + eyo_1 + eyo_2 + mutation_status + eyo_term_1*mutation_status + eyo_term_2*mutation_status + sex, data = data, family = gaussian(), prior = cauchy(), prior_intercept = cauchy(), chains = 8, cores = 1, iter = 10000, thin = 10)

**Table S2.**
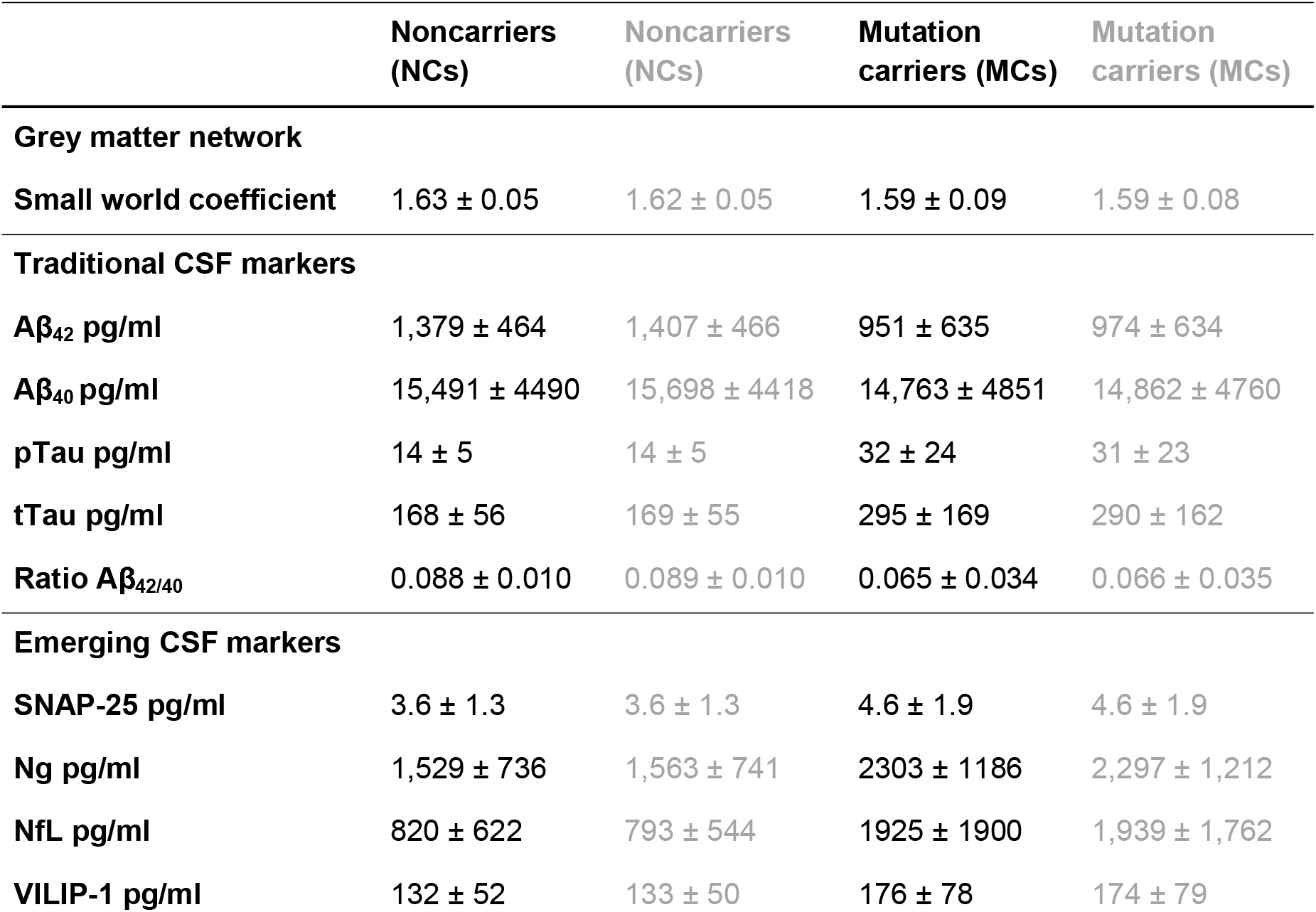
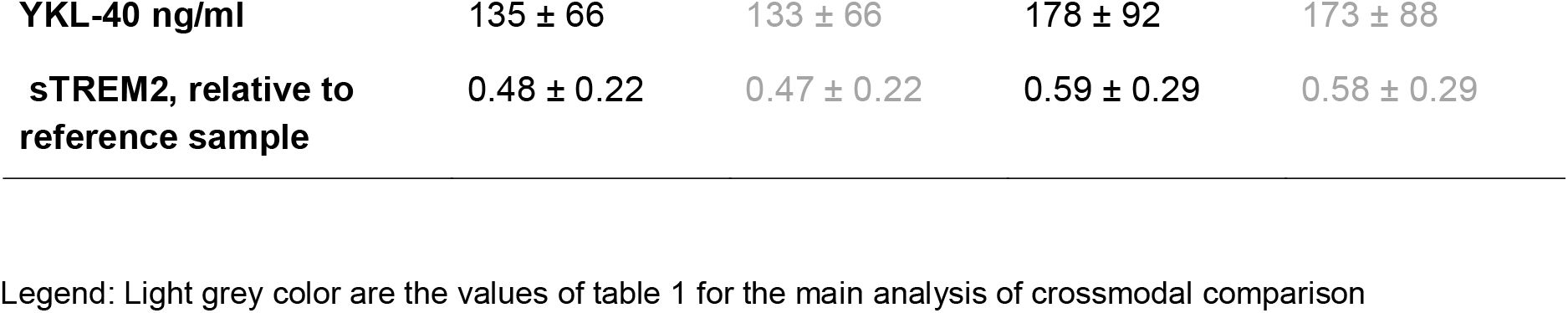
Baseline values for biomarkers used for EYO in comparison to crossmodal data

|  | **Noncarriers**<br>**(NCs)** | **Noncarriers**<br>**(NCs)** | **Mutation**<br>**carriers (MCs)** | **Mutation**<br>**carriers (MCs)** |
| --- | --- | --- | --- | --- |
| **Grey matter network** |  |  |  |  |
| **Small world coefficient** | 1.63 ± 0.05 | 1.62 ± 0.05 | 1.59 ± 0.09 | 1.59 ± 0.08 |
| **Traditional CSF markers** |  |  |  |  |
| **Aβ<sub>42</sub> pg/ml** | 1,379 ± 464 | 1,407 ± 466 | 951 ± 635 | 974 ± 634 |
| **Aβ<sub>40</sub> pg/ml** | 15,491 ± 4490 | 15,698 ± 4418 | 14,763 ± 4851 | 14,862 ± 4760 |
| **pTau pg/ml** | 14 ± 5 | 14 ± 5 | 32 ± 24 | 31 ± 23 |
| **tTau pg/ml** | 168 ± 56 | 169 ± 55 | 295 ± 169 | 290 ± 162 |
| **Ratio Aβ<sub>42/40</sub>** | 0.088 ± 0.010 | 0.089 ± 0.010 | 0.065 ± 0.034 | 0.066 ± 0.035 |
| **Emerging CSF markers** |  |  |  |  |
| **SNAP-25 pg/ml** | 3.6 ± 1.3 | 3.6 ± 1.3 | 4.6 ± 1.9 | 4.6 ± 1.9 |
| **Ng pg/ml** | 1,529 ± 736 | 1,563 ± 741 | 2303 ± 1186 | 2,297 ± 1,212 |
| **NfL pg/ml** | 820 ± 622 | 793 ± 544 | 1925 ± 1900 | 1,939 ± 1,762 |
| **VILIP-1 pg/ml** | 132 ± 52 | 133 ± 50 | 176 ± 78 | 174 ± 79 |
| **YKL-40 ng/ml** | 135 ± 66 | 133 ± 66 | 178 ± 92 | 173 ± 88 |
| **sTREM2, relative to**<br>**reference sample** | 0.48 ± 0.22 | 0.47 ± 0.22 | 0.59 ± 0.29 | 0.58 ± 0.29 |

Legend: Light grey color are the values of table 1 for the main analysis of crossmodal comparison

**Table S3.** Estimated years to onset of divergence between mutation carriers and noncarriers

|  | EYO of divergence<br>according 99%<br>credible interval | EYO of divergence<br>according 95%<br>credible interval | EYO of divergence,<br>according 99.5%<br>credible interval |
| --- | --- | --- | --- |
| **Grey matter network** |  |  |  |
| **Small world coefficient** | **-8** | **-10.4** | **-7.5** |
| **Traditional CSF markers** |  |  |  |
| **Aβ<sub>42/40</sub> ratio** | **-17.8** | **-18.4** | **-17.6** |
| **Aβ<sub>42</sub>** | **-15.5** | **-16.4** | **-15.2** |
| **Aβ<sub>40</sub>** | **0.5** | **-1** | **1.2** |
| **pTau** | **-17.7** | **-19.1** | **-17.2** |
| **tTau** | **-19.2** | **-20.4** | **-18.4** |
| **Emerging CSF markers** |  |  |  |
| **SNAP-25** | **-14.8** | **-17.9** | **-12.6** |
| **Ng** | **-19** | **-20.1** | **-18.2** |
| **NfL** | **-7** | **-8.1** | **-6.7** |
| **VILIP-1** | **-17.9** | **-19.8** | **-16.8** |
| **YKL-40** | **-7** | **-10.1** | **-6** |
| **sTREM2** | **-3.5** | **-6.7** | **-2.2** |

Legend: These analysis depend on sample sizes, which were for: small world N=439; Aβ <sub>42</sub>, Aβ <sub>40</sub>, pTau, & tTau N = 352; SNAP-25 & VILIP1 N=330, Ng & YKL-40 N=331, sTREM2 N=218; NfL N = 210.

**Figure S3.**
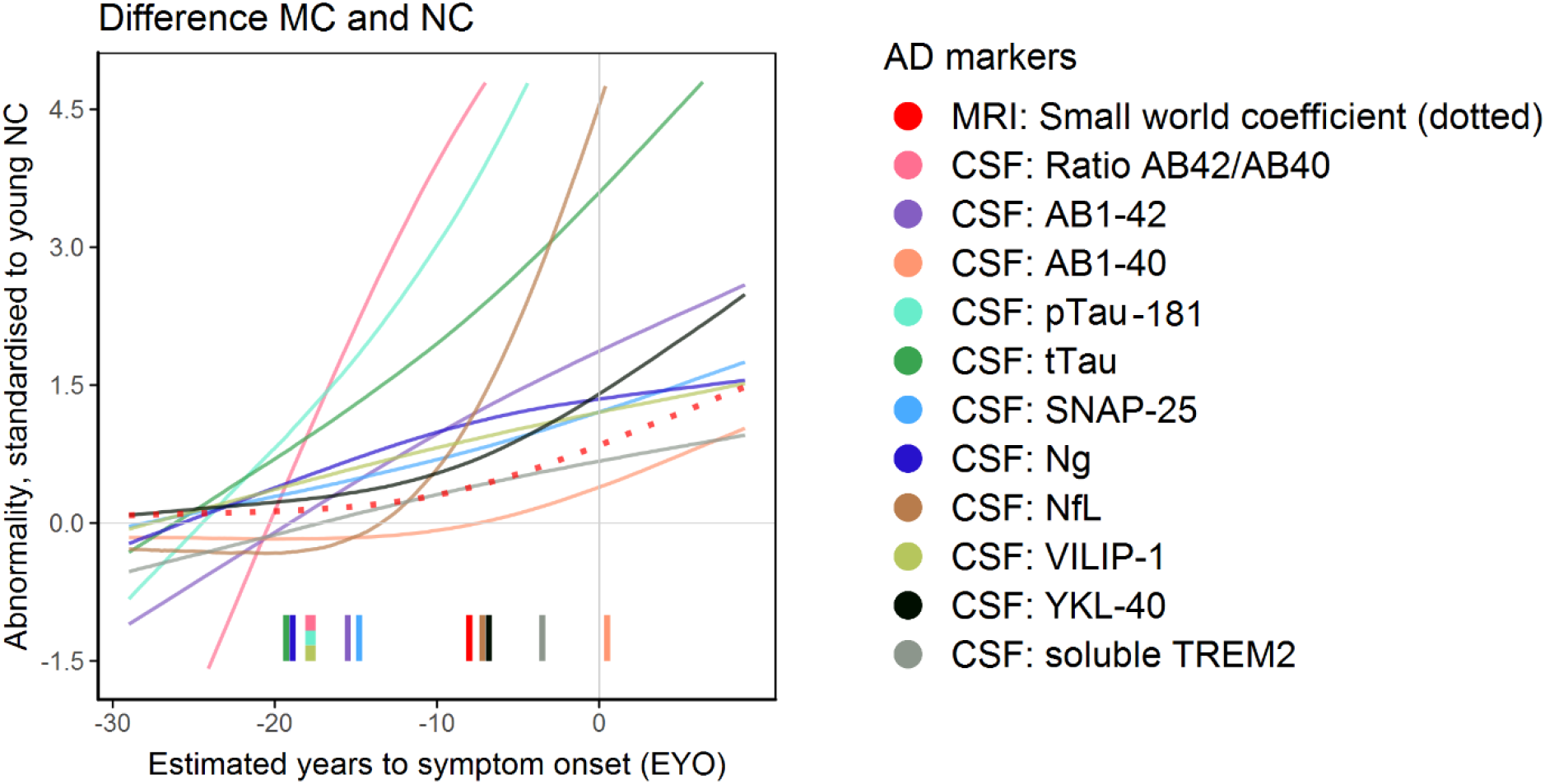
CSF and MRI biomarkers abnormality curves by EYO standardized to young noncarriers. Legend: The graphs show the median estimated curves standardized to the noncarrier mean and standard deviation (Table 1). All fitted lines are the median of the mixed models with a cubic spline, family random intercept and sex as covariate, and for the small world coefficient also total grey matter volume. These analyses depend on sample sizes, which were for: small world N=439; Aβ_42_, Aβ_40_, pTau, tTau N = 352; SNAP-25, VILIP1 N=330, Ng and YKL-40 N=331; sTREM2 N=218; NfL N=210. The tickmarks are the point that the 99% credible intervals of the difference between mutation carriers and noncarriers is different than 0 (for EYO divergence points at different thresholds, see Table S3)

